# Establishment of Reference Intervals for Peripheral Blood SII, NLR, PLR, and LMR in Children from the Zigong Region of China

**DOI:** 10.1101/2025.10.29.25339109

**Authors:** Xiaodan Zhang, Yaohui Song, Liangjun Zhang

## Abstract

**Objective:** To establish and validate reference intervals for the peripheral blood systemic immune-inflammation index (SII), neutrophil-to-lymphocyte ratio (NLR), platelet-to-lymphocyte ratio (PLR), and lymphocyte-to-monocyte ratio (LMR) in healthy children from the Zigong region of china.

**Methods:** A total of 2014 healthy children undergoing physical examination at The First People’s Hospital of Zigong from April 2023 to February 2025 were enrolled. Blood cell parameters were measured using the XN-1000 hematology analyzer, and SII, NLR, PLR, and LMR were calculated. The non-parametric method was employed to establish the 95% reference intervals.

**Results:** No statistically significant differences were observed in SII, NLR, PLR, or LMR between genders (P > 0.05). Participants were stratified into three age groups: 28 days to 2 years, 2 to 6 years, and 6 to 18 years. The established reference intervals for SII were 28.60×10^9^/L to 298.85×10^9^/L, 83.09×10^9^/L to 601.33×10^9^/L, and 140.02×10^9^/L to 657.19×10^9^/L, respectively, for the three age groups. The corresponding NLR reference intervals were 0.10-0.92, 0.35-1.88, and 0.57-2.30. PLR reference intervals were 29.39-115.15, 39.93-150.46, and 56.74-172.55. LMR reference intervals were 3.42-13.84, 2.81-13.03, and 2.41-10.56. A validation study conducted on 92 children from the Department of Child Health Care of the same hospital between April 2025 and September 2025 confirmed the applicability of these reference intervals. This indicates that the established reference intervals for peripheral blood SII, NLR, PLR, and LMR in children from the Zigong region are suitable for local clinical practice.

**Conclusion:** This study is the first to establish reference intervals for NLR, PLR, SII, and LMR in children from the Zigong region, providing a basis for the assessment of inflammatory diseases in local pediatric populations.

## 1. Introduction

In recent years, novel inflammatory biomarkers derived from routine blood tests - specifically, the systemic immune-inflammation index (SII), neutrophil-to-lymphocyte ratio (NLR), platelet-to-lymphocyte ratio (PLR), and lymphocyte-to-monocyte ratio (LMR) - have been increasingly applied in the diagnosis and prognosis of various diseases. Studies have demonstrated their significant value in conditions such as obesity, asthma, Kawasaki disease, infections, cancers, and autoimmune disorders ^1-7^.

While reference intervals for the aforementioned indices in adults and pregnant women have been widely reported ^8-10^, systematically established pediatric reference intervals remain scarce in China. The hematopoietic and immune systems in children undergo continuous development and dynamic maturation, resulting in normal hematological ranges that differ significantly from those of adults^11^. Direct application of local adult reference intervals may lead to misdiagnosis or delayed diagnosis in pediatric patients. This study aims to establish reference intervals for NLR, PLR, SII, and LMR in healthy children from the Zigong region.

## 2. Patients and Methods

### 2.1 Patients

From April 2023 to February 2025, a total of 2234 healthy children undergoing health examinations were initially enrolled in this study. The inclusion criterion was defined as healthy children aged 0-18 years from the Zigong region. Exclusion criteria were established in accordance with the guidelines outlined in “WS/T 779-2021 Reference Intervals for Child Blood Cell Analysis” ^12^, and included: (1) Diagnosis of congenital diseases; (2) Presence of fever or any acute diseases within the preceding two weeks; (3) History of chronic diseases, including hematological disorders (e.g., anemia, leukemia, platelet disorders), allergic conditions (e.g., eczema, urticaria, bronchial asthma), respiratory diseases (e.g., acute respiratory infections, lung malformations), urinary system diseases (e.g., Henoch-Schönlein purpura nephritis, glomerulonephritis, nephrotic syndrome), digestive system diseases (e.g., chronic diarrhea, inflammatory bowel disease), rheumatic immune diseases (e.g., rheumatoid arthritis, systemic lupus erythematosus), circulatory system diseases (e.g., myocarditis), endocrine and metabolic disorders (e.g., diabetes, thyroid diseases), malignancies, history of radiotherapy or chemotherapy, burns, or muscle trauma; (4) Underweight status, defined as a BMI-Z score less than -3 for children aged 5 years and below according to WHO growth standards, or a BMI below the age-specific screening threshold for underweight according to the Chinese industry standard WS/T 456 for children aged 6-18 years; (5) Recent medical history including: medication use (therapeutic drugs or supplements such as antibiotics, corticosteroids, or vitamin C) within one week; or history of surgery, blood transfusion, or significant blood loss within one month; (6) Abnormal laboratory findings: white blood cell count <3.0×10^9^/L or >15.0×10^9^/L, hemoglobin <90 g/L, or mean corpuscular volume (MCV) <75 fL. After applying the exclusion criteria, 2014 children (1015 males and 999 females) were included in the final analysis. Informed consent was obtained from the parents or legal guardians of all minor participants. The study protocol was reviewed and approved by the Ethics Committee of The First People’s Hospital of Zigong (NO.03202024).

### 2.2 Methods

Prior to blood collection, all participants maintained a healthy lifestyle for three days and fasted for 8-12 hours. Approximately 2 mL of peripheral venous blood was drawn into EDTA-K2 anticoagulant vacuum tubes and thoroughly mixed. All samples were analyzed within one hour of collection. Blood cell parameters, including platelet, monocyte, lymphocyte, and neutrophil counts, were determined using the Sysmex XN-1000 hematology analyzer (Sysmex Corporation, Japan) with its original reagents and quality controls. The SII, NLR, PLR, and LMR values were calculated using the following formulas: SII = (Platelet Count × Neutrophil Count) / Lymphocyte Count; NLR = Neutrophil Count / Lymphocyte Count; PLR = Platelet Count / Lymphocyte Count; LMR = Lymphocyte Count / Monocyte Count.

### 2.3 Statistical Analysis

Statistical analysis was performed using SPSS software (version 27.0). Extreme outliers were excluded based on predefined criteria: white blood cell count <3.0×10^9^/L or >15.0×10^9^/L, hemoglobin <90 g/L, or mean corpuscular volume (MCV) <75 fL. Further outlier identification was conducted using Tukey’s method, where the lower bound was defined as P_25_ - 1.5 × IQR and the upper bound as P_75_ + 1.5 × IQR; values outside this range were considered outliers and removed. The normality of data distribution was assessed using the Kolmogorov-Smirnov test. Normally distributed quantitative data are expressed as mean ± standard deviation (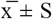), while non-normally distributed data are presented as median (P_25_, P_75_). In accordance with the CLSI EP28-A3C guidelines^13^, non-normally distributed data were subjected to Box-Cox transformation to approximate a normal distribution. For group comparisons, one-way ANOVA was used for comparisons across multiple groups, and the Mann-Whitney U test was applied for comparisons between two groups. A P-value < 0.05 was considered statistically significant. When a statistically significant difference was observed between groups, the Z-value and critical Z-value were compared to determine whether partitioning of reference intervals by sex and age was necessary. The Z-value was calculated as Z^*^ = 3 × √[(n_1_ + n_2_) / 240]. If Z > Z^*^, the between-group difference was considered clinically significant, warranting the establishment of separate reference intervals for the respective subgroups. The flow chart of this study is shown in Figure 1.

**Figure 1.**
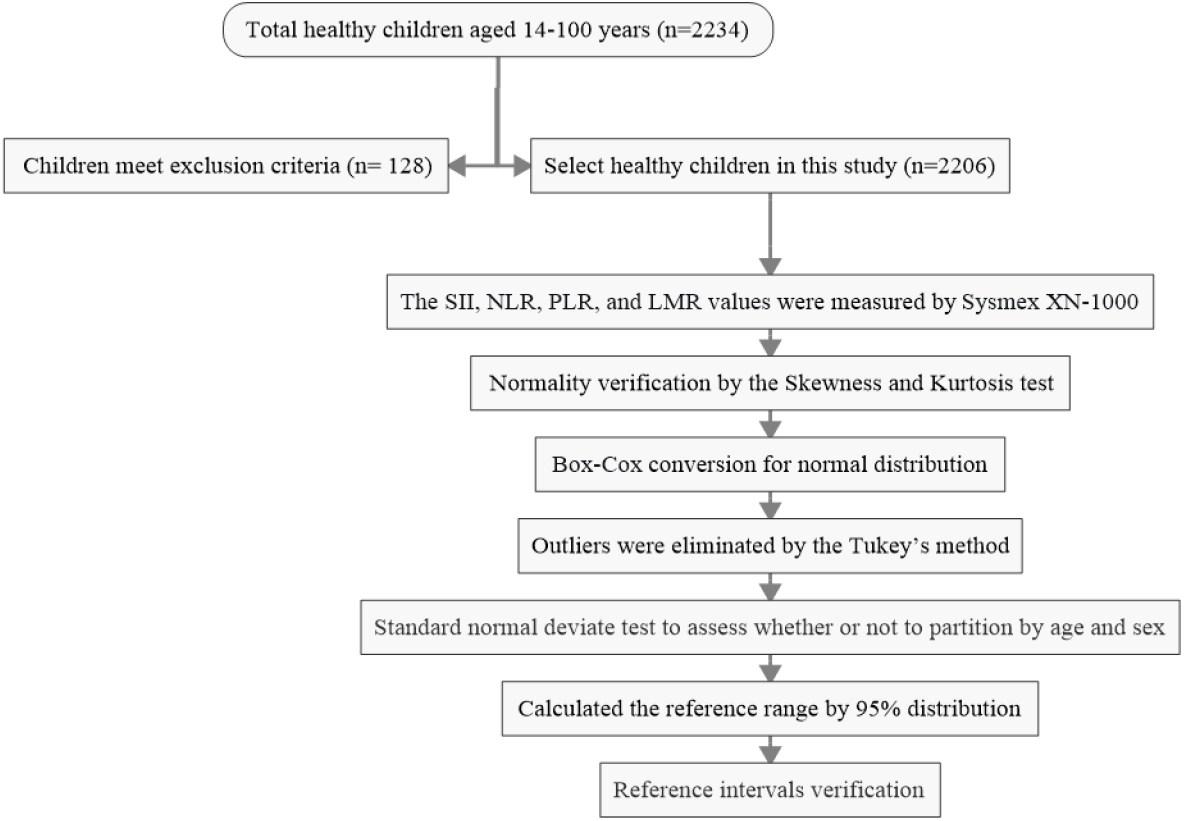
The flow chart of this study for establishing reference values.

## 3 Results

### 3.1 Distribution of data and elimination of outliers

This study followed the guidelines outlined in “WS/T 779-2021 Reference Intervals for Child Blood Cell Analysis” ^12^ issued by the National Health Commission of the People’s Republic of China. Participants were stratified into six age subgroups: 28 days to 6 months, 6 months to 1 year, 1-2 years, 2-6 years, 6-13 years, and 13-18 years. In accordance with the CLSI EP28-A3C guideline ^13^, reference intervals were established using a non-parametric approach. For skewed data distributions, the reference intervals were defined by the 2.5th to 97.5th percentiles^14^. The reference intervals for peripheral blood SII, NLR, PLR, and LMR were subsequently established and validated using this methodology. Based on the inclusion and exclusion criteria, 128 individuals were excluded. An additional 192 outliers were identified and removed using Tukey’s method, resulting in a final cohort of 2014 children (1015 males and 999 females). (Table 1)

**Table 1.** Distribution of the 2014 Healthy Children by Sex and Age.

| Age | n | male | female |
| --- | --- | --- | --- |
| 28 days-6 months | 154 | 76 | 78 |
| 6 months-1 year | 159 | 84 | 75 |
| 1-2 years | 216 | 101 | 115 |
| 2-6 years | 419 | 211 | 208 |
| 6-13 years | 860 | 442 | 418 |
| 13-18 years | 206 | 101 | 105 |

### 3.2 Normality Test and Transformation

Normality of the peripheral blood indices (SII, NLR, PLR, and LMR) was assessed using the Kolmogorov-Smirnov test. The results indicated that none of the parameters followed a normal distribution (P < 0.05). Subsequently, the data were transformed using the Box-Cox method. Upon re-evaluation, the transformed data showed no significant deviation from normality (P > 0.05) (Table 2).

**Table 2.** Normality Test Results for SII, NLR, PLR, and LMR Before and After Transformation.

| Parameter | Before Transformation |  |  | After Transformation |  |  |
| --- | --- | --- | --- | --- | --- | --- |
|  | Skewness | Kurtosis | K-S Asymptotic Significance | Skewness | Kurtosis | K-S Asymptotic Significance |
| SII | 0.023 | -0.275 | <0.001 | 0.000 | -0.028 | 0.972 |
| NLR | 0.006 | -0.310 | <0.001 | 0.000 | -0.029 | 0.950 |
| PLR | 0.040 | -0.278 | <0.001 | 0.000 | -0.028 | 1.000 |
| LMR | 0.035 | -0.051 | <0.001 | 0.016 | -0.067 | 0.192 |

### 3.3 Comparison of SII, NLR, PLR, and LMR by Sex

Although the comparisons of SII, NLR, PLR, and LMR between males and females showed statistically significant differences (P < 0.05) in their concentration levels, the Z-test results indicated that the Z-values for all these parameters were lower than the critical Z^*^-value. This suggests that the observed sex-based differences are unlikely to be of clinical relevance. Furthermore, no statistically significant difference was found for LMR between sexes (P > 0.05) (Table 3). Consequently, establishing separate reference intervals by gender was deemed unnecessary for all parameters.

**Table 3.** Comparison of SII, NLR, PLR, and LMR Levels Between Males and Females.

| Parameter | T value | P value | Z -test |  |
| --- | --- | --- | --- | --- |
|  |  |  | Z value | Z* value |
| SII | 3.411 | 0.000 | 3.411 | 8.392 |
| NLR | 2.838 | 0.005 | 2.838 | 8.392 |
| PLR | 2.403 | 0.016 | 2.403 | 8.392 |
| LMR | 1.950 | 0.051 | - | - |

### 2.4 Comparison of SII, NLR, PLR, and LMR Across Age Groups

The reference population was stratified by age. One-way ANOVA revealed that the differences in NLR levels between the following adjacent age groups were statistically significant (P < 0.05): 6 months-1 year vs. 1-2 years, 1-2 years vs. 2-6 years, and 2-6 years vs. 6-13 years. Similarly, significant differences (P < 0.05) were observed for SII, PLR, and LMR between the 1-2 years vs. 2-6 years and 2-6 years vs. 6-13 years age groups. However, subsequent Z-test analysis showed that only for NLR between the 6 months-1 year and 1-2 years groups was the Z-value less than the critical Z^*^-value, indicating that this specific difference was not clinically significant and did not warrant separate reference intervals. For all other parameters and comparisons where no statistically significant difference was found (P > 0.05), partitioning was also deemed unnecessary (Table 4). Based on these results, reference intervals for SII, NLR, PLR, and LMR were established for three consolidated age groups: Group 1 (28 days-2 years, combining the original 28 days-6 months, 6 months-1 year, and 1-2 years subgroups), Group 2 (2-6 years), and Group 3 (6-18 years, combining the original 6-13 years and 13-18 years subgroups).

**Table 4.**
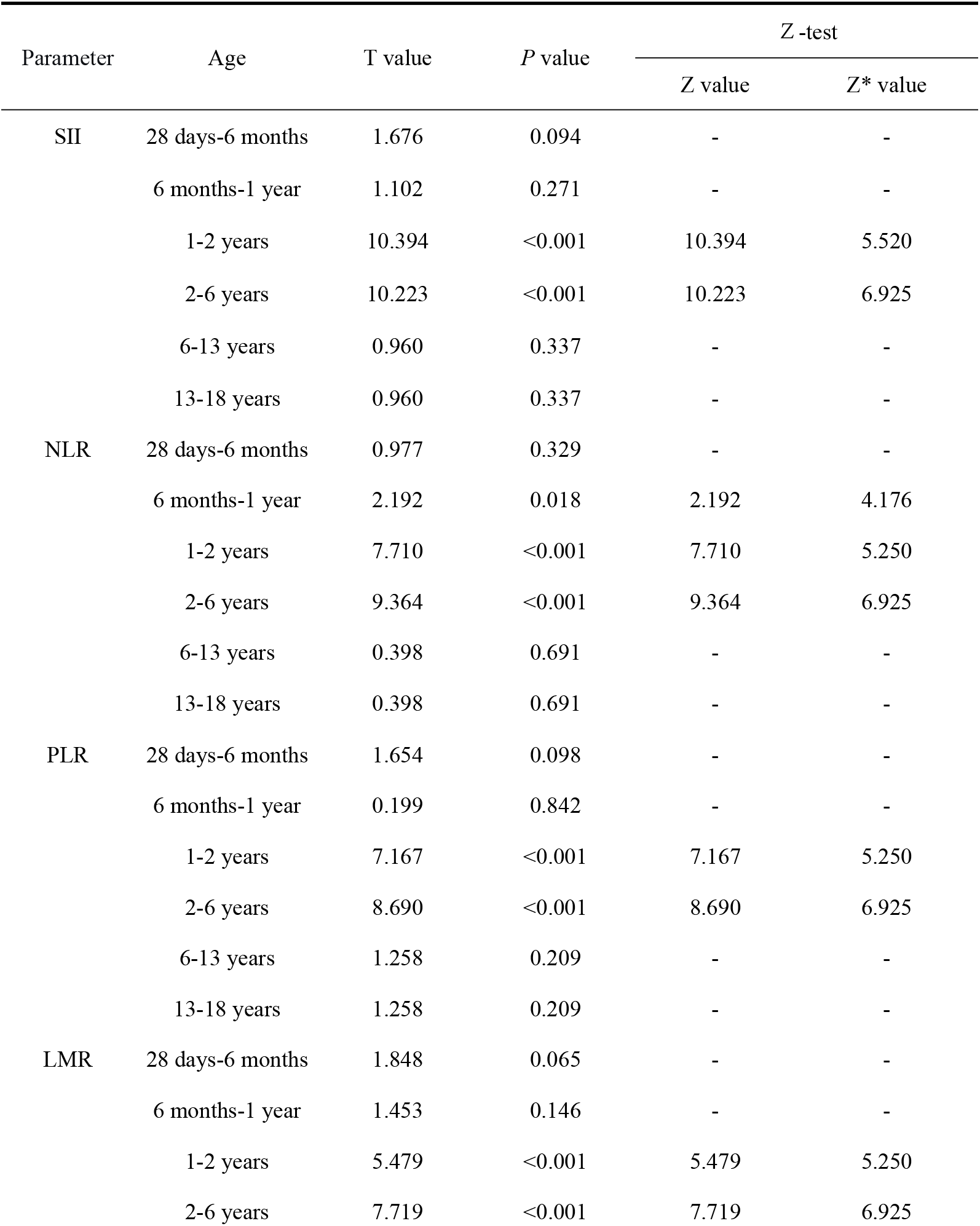
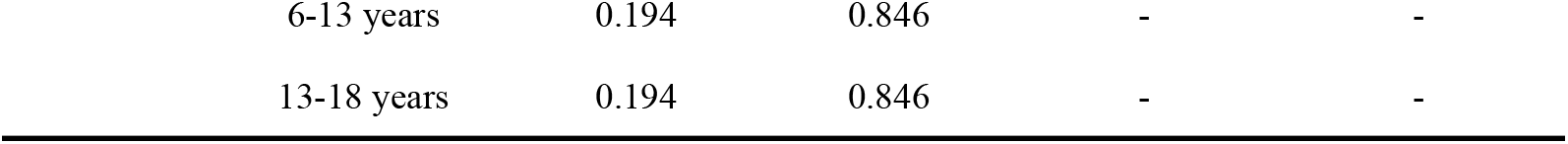
Comparison of SII, NLR, PLR, and LMR Levels Across Age Groups.

### 3.5 Established Reference Intervals for SII, NLR, PLR, and LMR

Reference intervals for each parameter were established by the nonparametric method for the respective age groups using the percentile method (P_2.5_ to P_97.5_) and are presented in Table 5. Analysis of the intervals revealed that SII, NLR, and PLR levels tended to increase with age, whereas LMR demonstrated a decreasing trend (Figure 2).

**Table 5.** The SII, NLR, PLR, and LMR reference interval calculate by nonparametric methods.

| Parameter | n | Age | Reference intervals |
| --- | --- | --- | --- |
| SII($\times 10^9/L$) | 529 | 28 days-2 years | 99.88 (28.60-298.85) |
|  | 419 | 2-6 years | 229.71 (83.09-601.33) |
|  | 1066 | 6-18 years | 328.71 (140.02-657.19) |
| NLR | 529 | 28 days-2 years | 0.30 (0.10-0.92) |
|  | 419 | 2-6 years | 0.81 (0.35-1.88) |
|  | 1066 | 6-18 years | 1.19 (0.57-2.30) |
| PLR | 529 | 28 days-2 years | 62.58 (29.39-115.15) |
|  | 419 | 2-6 years | 83.98 (39.93-150.46) |
|  | 1066 | 6-18 years | 103.79 (56.74-172.55) |
| LMR | 529 | 28 days-2 years | 8.97 (3.42-13.84) |
|  | 419 | 2-6 years | 7.00 (2.81-13.03) |
|  | 1066 | 6-18 years | 5.86 (2.41-10.56) |

**Figure 2.**
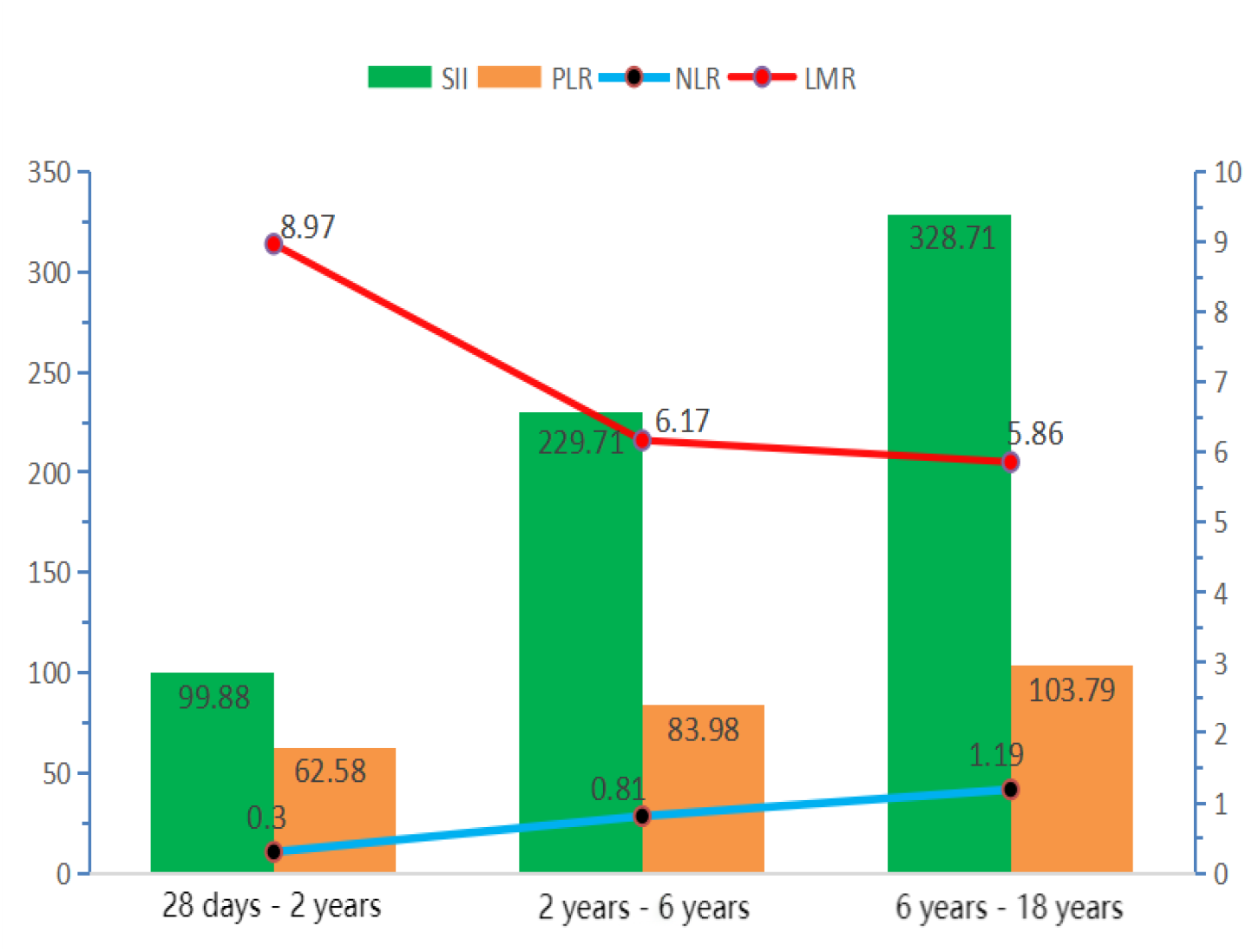
Trends in SII, NLR, PLR, and LMR Across Age Groups

### 3.6 Verification of Reference Intervals

In accordance with WS/T 402 ^11^, the established reference intervals were verified using the “healthy population sampling verification method.” A total of 92 children (47 males and 45 females) undergoing health examinations in the Department of Child Health at the First People’s Hospital of Zigong from April to September 2025 were selected for validation. The age distribution was as follows: 31 in the 28 days-2 years group, 29 in the 2-6 years group, and 32 in the 6-18 years group. All validation samples met the inclusion criteria outlined in section 1.1 and shared consistent characteristics with the original cohort used to establish the reference intervals. Following the CLSI EP28-A3C guideline^13^, the verification was considered successful if the proportion of validation results falling outside the reference intervals was ≤10%. Our results confirmed that the established reference intervals for peripheral blood SII, NLR, PLR, and LMR in children from the Zigong region are suitable and applicable for local clinical practice.

### 3.7 Comparison of SII, NLR, PLR, and LMR Reference Intervals Across Different Regions in China

This study compared the established reference intervals with those reported from Guangdong ^15^, Shenyang ^16^, Hebei ^17^, and Shenzhen^18^ in china (Table 6). The results revealed notable differences in the reference intervals for SII, NLR, PLR, and LMR among children of the same age groups across these regions in china. Specifically, the SII reference intervals in the present study demonstrated lower lower limits and higher upper limits compared to those from Guangdong, Shenyang, and Shenzhen for corresponding age groups. In contrast, the upper limit of the SII reference interval reported in Hebei was 130 units higher than that of the present study for the same age group. For NLR, PLR, and LMR, the reference intervals established in this study exhibited lower lower limits and higher upper limits relative to those from Guangdong Shenyang, Hebei, and Shenzhen in the same age categories.

**Table 6.** Comparison of Reference Intervals Across Different Regions in China.

| Region | Age | n | Reference Intervals |  |  |  |
| --- | --- | --- | --- | --- | --- | --- |
| | | | SII($\times 10^9/L$) | NLR | PLR | LMR |
| Guangdong<sup>15</sup> | 28 days-2 years | 198 | 187.4<br>(109.8,219.9) | 0.58<br>(0.33,0.68) | 69.9 (48.7,78.5) | - |
| Shenyang<sup>16</sup> | 2-6 years | 60 | 179.86<br>(101.27,210.95) | 0.55<br>(0.35,0.83) | 72.58<br>(59.10,82.7) | - |
| Hebei<sup>17</sup> | 2-6 years | 50 | 640. 43±105. 43 | 1. 05±0. 51 | - | - |
| Shenzhen<sup>18</sup> | 6-18 years | 150 | 325.75<br>(231.51,437.59) | 1.07<br>(0.73,1.34) | 109.32<br>(86.02,126.19) | 6.67<br>(5.26,8.33) |

## 4. Discussion

This study is the first to systematically establish age-specific reference intervals for SII, NLR, PLR, and LMR in healthy children aged 28 days to 18 years in the Zigong region of China. Our results indicate no clinically significant sex-based differences for these parameters; however, they exhibited clear age-dependent trends: SII, NLR, and PLR gradually increased with age, while LMR demonstrated a progressive decline. These trends align closely with established patterns of hematopoietic development in children.

During infancy, lymphocytes constitute up to 60% of circulating leukocytes. Neutrophil levels gradually increase during the preschool years, with the neutrophil-to-lymphocyte ratio reaching 1:1 by 4-6 years of age. After age 6, neutrophils become more predominant while lymphocytes decrease, progressively approaching adult proportions ^19^. Accordingly, we observed that NLR increased with age, approaching a value of 1 in the 2-6 years group, consistent with this developmental process. PLR also showed a general increasing trend, though it was less pronounced, which can be attributed to the wide reference intervals for platelets throughout childhood ^12^, even when considering the unique developmental trajectory of lymphocytes ^19^.

LMR displayed a distinct decline with age, particularly marked between the 28 days-2 years and 2-6 years groups. This sharp decrease corresponds to the period when lymphocytes can reach up to 60% around 2-3 years of age ^19^, followed by a gradual decline, while monocyte counts remain relatively stable ^12^. The decline in LMR was less pronounced from the 2-6 years to the 6-18 years group, consistent with the stabilization of lymphocyte percentages at 20%-40% after approximately 7 years of age ^18^. These dynamic changes in SII, NLR, PLR, and LMR are intrinsically linked to the maturation of the pediatric immune system, where ongoing development of immune organs and evolving cellular populations and functions lead to age-associated variations in these inflammatory indices ^20^. These physiological changes underscore the necessity of establishing age-specific reference intervals for children, as the direct application of adult ranges may lead to clinical misinterpretation.

Comparisons with studies from other regions in China ^15-18^ revealed distinct differences in the established reference intervals. Specifically, the SII intervals in our study had lower lower limits and higher upper limits than those reported for corresponding age groups in Guangdong, Shenyang, and Shenzhen, whereas the upper limit for SII in Hebei was 130 units higher than our value. Similarly, our reference intervals for NLR, PLR, and LMR generally demonstrated lower lower limits and higher upper limits compared to other regions. Several factors may explain these regional variations. First, differences in sample size may contribute ^20^. Our study featured a larger total sample size (n=2014) with each age subgroup containing more than 120 individuals, adhering to CLSI recommendations ^13^, which likely yields more robust estimates compared to the smaller samples in other studies ^15-18^. Second, regional environmental factors-such as climate, humidity, air quality, and sunlight exposure-may influence respiratory mucosal barrier integrity and immune cell activity, potentially causing fluctuations in these inflammatory markers ^21^. Genetic polymorphisms across different populations could also affect the efficiency of immune cell differentiation and maturation, leading to inherent differences in immunological parameters ^22^. Furthermore, regional variations in dietary patterns and hygiene practices ^23-24^ may modulate immune system homeostasis, thereby influencing the established reference intervals. These comparative findings reinforce the necessity of establishing region-specific reference intervals for laboratory parameters.

## 5. Conclusion

In conclusion, this study successfully establishes reference intervals for SII, NLR, PLR, and LMR in children from the Zigong region, providing a critical foundation for local pediatric clinical practice. Future multicenter studies are warranted to further explore the diagnostic and prognostic utility of these inflammatory indices in specific childhood diseases.

## Data Availability Statement

The data supporting the findings of this study are available from the corresponding author upon reasonable request.

## Ethical approval

This study was approved by the Ethics Committee of the Zigong First People’s Hospital (NO.03202024).

## Author Contributions

Conceptualization, L.Z.; methodology, L.Z. and X.Z.; formal analysis, L.Z. and X.Z; investigation, X.Z.; writing-original draft preparation, X.Z.; writing-review and editing, X.Z.; visualization, Y.S.; supervision, Y.S. and L.Z.; project administration, Y.S. and L.Z. All authors have read and agreed to the published version of the manuscript.

## Acknowledgements

None

